# Detecting change-points in preclinical rheumatoid arthritis biomarkers using Bayesian multivariate segmented regression

**DOI:** 10.64898/2026.05.22.26353892

**Authors:** Yonatan F. Wolde, Alexandria M. Jensen, Brandie D. Wagner, Jess D. Edison, Marie L. Feser, Michael Mahler, Kevin D. Deane, Kevin P. Josey

## Abstract

**Background:** Rheumatoid arthritis (RA) has a preclinical period characterized by elevations in serum autoantibodies. Identifying the timing and magnitude of autoantibody trajectory changes may inform screening strategies and preventative interventions.

**Methods:** Using a Bayesian multivariate segmented regression, we jointly modeled longitudinal autoantibody trajectories from two Department of Defense Serum Repository cohorts (Cohort A: 209 matched case-control pairs, 1566 samples, six autoantibody biomarkers; Cohort B: 309 cases with two matched controls each, 2758 samples, eight autoantibody biomarkers). Change-points and magnitudes of change were estimated simultaneously under a multivariate normal likelihood on log-transformed concentrations, treating values at each assay’s upper limit of detection as right-censored through data augmentation and modeling an unstructured residual correlation among biomarkers; a diagonal-covariance sensitivity analysis is reported in the Supplement.

**Results:** In Cohort A, 5/6 biomarkers shifted before diagnosis, with 95% highest posterior density intervals (HPDIs) excluding zero. RF-IgA changed earliest, 9.2 years before diagnosis (HPDI: −13.8, −4.2), followed by RF-IgM at 8.2 years (−11.7, −5.4) and ACPA-IgG at 7.8 years (−9.9, −6.1). In Cohort B, all eight biomarkers changed before diagnosis, anti-CCP3 (IgG) earliest at 7.2 years (−8.1, −6.4); IgG isotypes changed earlier and more steeply than IgA. Ranked by a composite time integrating timing and magnitude, ACPA-IgG and anti-CCP3 (IgG) came first.

**Conclusions:** This Bayesian model estimates change-points and magnitudes simultaneously while accounting for the upper limits of detection and for correlation among biomarkers, and it characterizes the uncertainty in both. It complements prior divergence-based methods for understanding preclinical RA autoimmunity.

**Key Messages:**

- Treating autoantibody concentrations at the upper limit of detection as right-censored and modeling the residual correlation among biomarkers allow joint estimation of when and how steeply each autoantibody begins to rise before RA diagnosis.
- RF-IgA, RF-IgM, and ACPA-IgG exhibited the earliest pre-diagnostic trajectory acceleration (7.8 to 9.2 years before diagnosis), and all eight markers in a second panel of anti-citrullinated protein antibodies shifted before diagnosis, with IgG isotypes changing earlier than IgA.
- A composite time integrating the timing and magnitude of autoantibody change provides a different ranking of biomarkers than change-point estimation alone, which may inform future screening and prevention strategies.

## Introduction

Rheumatoid arthritis (RA) is an autoimmune disorder that affects 0.5-1% of the population.^1,2^ The disease imposes considerable burden through systemic inflammation and joint damage, making early identification and intervention an important public health priority. Recent findings in RA pathogenesis have revealed a well-characterized preclinical stage that precedes clinical RA by years or even decades, presenting opportunities for early disease detection and prevention strategies.^3^

This preclinical RA stage is characterized by progressive development of autoimmune responses that can be detected by the emergence and evolution of serum autoantibodies.^4–8^ Multiple autoantibodies have been identified in preclinical RA, including biomarkers that are well-established in RA such as rheumatoid factor (RF) and anti-citrullinated protein antibodies (ACPA). ACPA is a general class of autoantibodies, and the version used most often in clinical care is the anti-cyclic citrullinated peptide (anti-CCP) antibody. Both RF and ACPA can be of a variety of isotypes including immunoglobulin (Ig) A, IgM and IgG. Emerging data show that other autoantibodies, including antibodies to specific citrullinated proteins and to isoenzymes of peptidyl arginine deiminase (anti-PAD), can be present in preclinical RA.^9–11^

Characterizing the sequence and timing of autoantibody emergence informs how autoimmune responses progress, and identifying which autoantibodies appear earliest and change most sharply helps researchers develop biomarker-based screening strategies for at-risk populations.^12^ These temporal patterns may also inform when clinical RA is likely to develop and when preventive treatment might be offered, an active area of investigation given recent trials showing that onset can be delayed or prevented in high-risk individuals.^3,13^

Previous investigations focusing on preclinical RA autoantibody trajectories have advanced the field, yet they have methodological limitations. In particular, using a longitudinal dataset derived from the Department of Defense Serum Repository (DoDSR), Kelmenson and colleagues^14^ modeled six autoantibodies in preclinical RA using mixed-effects cubic B-splines. These analyses identified an approximate change-point of each autoantibody by selecting the earliest time at which the cubic function of cases and controls began to diverge. However, they did not account for the upper limit of detection (ULoD) in these assays, fully characterize the uncertainty of the change-point, or formally incorporate the magnitude of autoantibody elevation. Several autoantibodies exceed the ULoD close to disease onset; treating those capped measurements as exact, or removing them through truncation, biases both the timing and the magnitude of the estimated change.

We investigate when each autoantibody’s trajectory changes in the preclinical stage of RA to determine which biomarkers show the earliest and most pronounced changes in serum concentrations. Our Bayesian multivariate segmented regression treats concentrations at the ULoD as right-censored, models the residual correlation among biomarkers, characterizes uncertainty through posterior distributions, and incorporates both the timing and magnitude of change in the biomarker’s serum concentration. We apply it to two autoantibody biomarker panels. The first contains six autoantibodies previously studied in this DoDSR cohort, enabling direct comparison with prior findings. The second contains eight autoantibodies measured with multiplex technology, which have previously been examined in this cohort only as positive or negative results.^9^

## Methods

### Data Collection and Study Design

We used a cohort of individuals who developed clinical RA and matched controls obtained from the DoDSR.^9,14^ In brief, this cohort includes longitudinal clinical data and serum samples obtained prior to and after a diagnosis of clinical RA based on established classification criteria^15^ or diagnosis by a board-certified rheumatologist from individuals who developed RA while on active-duty military service.

We evaluated two subsets of subjects from this larger cohort: Cohort A, which included cases matched to a single control; and Cohort B, which included cases that have two matched controls. Controls were matched to cases based on sex, race, age, and region of enlistment, and were selected to have a similar longitudinal timespan to that of their matched case. In each subset, only observations with complete biomarker data were included. A summary of the final datasets is described in Table 1.

**Table 1:** Characteristics of Cohort A and Cohort B among participants who develop (RA) and their matched counterparts who do not develop (non-RA) rheumatoid arthritis.

|  |  | Cohort A (209 pairs; 1:1 match) |  | Cohort B (309 pairs; 1:2 match) |  |
| --- | --- | --- | --- | --- | --- |
|  |  | Non-RA | RA | Non-RA | RA |
| **Sex** | Male | 108 (51.7%) | 108 (51.7%) | 306 (49.5%) | 153 (49.5%) |
|  | Female | 101 (48.3%) | 101 (48.3%) | 312 (50.5%) | 156 (50.5%) |
| **Race** | White | 121 (57.9%) | 121 (57.9%) | 326 (52.8%) | 163 (52.8%) |
|  | Black | 55 (26.3%) | 55 (26.3%) | 158 (25.6%) | 79 (25.6%) |
|  | Hispanic/Latino | 18 (8.6%) | 18 (8.6%) | 74 (12.0%) | 37 (12.0%) |
|  | Other | 15 (7.2%) | 15 (7.2%) | 60 (9.7%) | 30 (9.7%) |
| **Age at diagnosis** | Median (IQR) | 36.3 (30.6-42.4) | 36.2 (30.8-42.1) | 34.9 (28.7-40.6) | 34.7 (29.0-40.5) |
| **Number of serum measures** | N (Average) | 734 (3.5) | 832 (4.0) | 1798 (2.9) | 960 (3.1) |

Variables were measured at enrollment and do not vary over time within participants.

In Cohort A, which is the same set used in the publication by Kelmenson and colleagues,^14^ we evaluated six autoantibody biomarkers: ACPA IgA, IgG, and IgM, and RF IgG, IgA, and IgM. These assays were performed on an enzyme-linked immunosorbent assay (ELISA; Inova Diagnostics, Inc., San Diego, California). In Cohort B, we evaluated eight autoantibody biomarkers, comprising both IgG and IgA isotypes of antibodies to cyclic citrullinated peptide-3 (anti-CCP3, the target of which contains a proprietary mix of citrullinated antigens), and antibodies to the following specific citrullinated proteins: citrullinated vimentin-2 (anti-citVim2), cit-fibrinogen (anti-citFib) and citrullinated histone-1 (anti-citHis1). These eight biomarkers were evaluated on a fully automated particle-based multianalyte technology (Inova Diagnostics, Inc.) and the Aptiva system, which uses paramagnetic particles with unique signatures and a digital interpretation system, as described previously.^16^ These eight biomarkers were selected because, in prior analyses using this same cohort and using dichotomous cut-offs for each marker, their IgG isotypes were found to be positive at meaningfully higher rates in RA cases compared to controls.^9^

### Statistical Analysis

We employed a Bayesian segmented linear mixed modeling approach to jointly analyze serum biomarker trajectories over time.^17–19^ We analyzed the two cohorts separately. The primary response for each observation was the vector of log-transformed serum biomarker measurements, which we assume follows a multivariate normal distribution, right-censored at each assay’s upper limit of detection. The log transformation stabilizes the right-skewed assay distributions and makes the normal residual model plausible for strictly positive concentrations. Each post-change slope is then an additive change in the slope of log concentration, which corresponds to a multiplicative change in concentration per year. In Cohort B, seven measurements were recorded as exactly zero; we set each to half the smallest positive value observed for that biomarker before taking logs. For participant *i* = 1,2, …, *n*, measurement within a participant *j* = 1,2, …, *m_i_*, *l* = 1,2, …, *p* denoting case-control pairs (we use *l*[*i*] to identify patient pairs corresponding with participant *i*), and for biomarker *k* = 1, 2, …, *K* (*K* = 6 for Cohort A; *K* = 8 for Cohort B), the multivariate outcomes are denoted with ***Y****_i_*_j_ = [*Y_i_*_j1_, *Y_i_*_j2_, …, *Y_i_*_j*K*_]^T^ with each element *Y_ijk_* denoting the log concentration of the corresponding biomarker. Each raw concentration is recorded up to the *k*^th^ biomarker’s maximum detectable value; on the log scale we denote these limits as ***U*** = [*U*_1_, *U*_2_, …, *U_K_*]^T^. The response distribution is

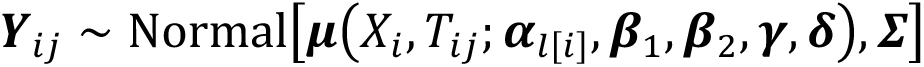

where ***Σ*** is a residual variance-covariance matrix. Values recorded at the upper detection limit are right-censored: the assay states only that the true concentration lies at or above the limit. We handle censoring with data augmentation.^20,21^ Each censored measurement is replaced by a latent value constrained to lie at or above the limit. The latent values are sampled jointly with the model parameters, and the completed outcome vectors follow the multivariate normal likelihood above, which preserves the residual correlation between censored and observed components of the same vector. Integrating over the latent values recovers the censored-data likelihood exactly. Modeling an unstructured residual correlation lets the biomarkers share information.^22–24^ The mean structure is defined as:

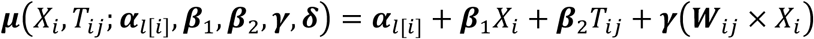

where *X_i_* ∈ {0,1} is an indicator for whether the participant is diagnosed with RA*, T_i_*_j_ is the time before diagnosis of the case in the case-control pair, and ***W****_i_*_j_ = pmax{**0**, *T_i_*_j_ – ***δ***} with ***δ*** = (*δ*_1_, *δ*_2_, …, *δ_K_*)^T^ representing the biomarker-specific change-points. The paired random intercept ***α****_l_* ∼ N(***θ***, ***Ω***), where ***Ω*** = diag(***ω***^2^) is the diagonal matrix consisting of entries ***ω*** = (*ω*_1_, *ω*_2_, …, *ω_K_*)^T^, accounts for the correlation among repeated measurements within a matched set^22,23^ and captures variation due to time-invariant characteristics (e.g., sex, race, and age at diagnosis). ***γ*** quantifies the magnitude of change (the change in slope) among RA cases. The residual covariance matrix Z is parametrized as Z = diag(***σ***^2^) ⊙ ***Φ*** where ***Φ*** is a (*K* × *K*) correlation matrix and diag(***σ***^2^) is the diagonal matrix of measurement variances with ***σ*** = (*σ*_1_, *σ*_2_, …, *σ_K_*)^T^.

Weakly informative priors, the distributions assigned to parameters before the data are observed, were placed on all regression parameters and hyperparameters^25,26^: mean-zero normal priors with variance = 4 on the slope parameters (on the log scale), diffuse mean-zero normal priors on the intercepts, half-Cauchy priors on the standard deviations, and a uniform prior over [−20, 10] years on each change-point. The correlation matrix ***Φ*** received an LKJ(2) prior on its Cholesky factor, which is weakly informative and centered on the identity. The Supplement lists each prior in full.

In addition to the change-points *δ_k_*, and post-change slopes *γ_k_*, we also summarize both jointly through a composite time *τ_k_*, defined as the earliest time satisfying Pr{*γ_k_*(*τ_k_* − *δ_k_*) > 0 ∣ *X_i_* = 1} = 0.9. This is the point at which there is high posterior probability that the biomarker’s mean among cases has deviated from its baseline trend. Biomarkers in Table 2 are ranked by *τ_k_*. A sensitivity analysis that fixes the residual correlation to the identity, leaving every other model component unchanged, is reported in the Supplement.

**Table 2:** Composite time, average change-point, and magnitude of change for each biomarker among RA cases, ordered by composite time within each cohort.

| Cohort A |  |  |  |
| --- | --- | --- | --- |
| Biomarker | Composite Time (Years) | Average Change-point [95% HPDI] | Magnitude of Change Mean [95% HPDI] |
| ACPA IgG | -6.6 | -7.8 [-9.9, -6.1] | 0.25 [0.20, 0.31] |
| RF IgM | -6.4 | -8.2 [-11.7, -5.4] | 0.13 [0.09, 0.17] |
| RF IgA | -6.0 | -9.2 [-13.8, -4.2] | 0.11 [0.07, 0.16] |
| ACPA IgA | -4.5 | -6.1 [-8.9, -3.7] | 0.13 [0.09, 0.18] |
| ACPA IgM | -3.4 | -5.2 [-8.6, -2.4] | 0.06 [0.03, 0.10] |
| RF IgG | 4.6 | 4.0 [2.6, 4.9] | 2.52 [0.50, 4.85] |
| Cohort B |  |  |  |
| anti-CCP3 (IgG) | -6.7 | -7.2 [-8.1, -6.4] | 0.36 [0.32, 0.41] |
| anti-citHis1 (IgG) | -5.9 | -7.0 [-8.5, -5.5] | 0.17 [0.14, 0.21] |
| anti-citVim2 (IgG) | -5.8 | -6.5 [-7.5, -5.5] | 0.22 [0.19, 0.26] |
| anti-citFib (IgG) | -5.1 | -5.9 [-7.1, -4.7] | 0.18 [0.15, 0.21] |
| anti-citFib (IgA) | -4.3 | -5.8 [-8.4, -3.5] | 0.04 [0.02, 0.06] |
| anti-citVim2 (IgA) | -3.9 | -5.7 [-8.0, -2.9] | 0.05 [0.03, 0.07] |
| anti-citHis1 (IgA) | -3.8 | -5.7 [-9.0, -2.9] | 0.06 [0.03, 0.08] |
| anti-CCP3 (IgA) | -3.7 | -4.5 [-5.7, -3.2] | 0.12 [0.09, 0.15] |

Change-point estimates represent the time (in years before diagnosis) when biomarker trajectories begin to diverge from baseline trend among RA cases. The 95% highest posterior density intervals (HPDI) reflect uncertainty in change-point timing. Composite Time denotes the earliest time point (in years relative to RA diagnosis) at which there is 90% posterior probability that the mean among cases has departed from its baseline trend. Negative values indicate pre-diagnostic change-points; positive values indicate post-diagnostic change-points. Magnitudes of change are slopes of log concentration per year. **Abbreviations:** RF=rheumatoid factor; Ig=immunoglobulin; ACPA=anti-citrullinated protein antibody; CCP3=cyclic citrullinated peptide-3; cit=citrullinated; His=histone; Vim=vimentin; Fib=fibrinogen; HPDI=highest posterior density interval.

Posterior inference was conducted using Markov Chain Monte Carlo (MCMC) sampling in Stan.^27^ Five chains, each containing 20 000 iterations, were simulated, with the first 10 000 discarded as a warm-up and a thinning interval of ten applied. We assessed mixing with diagnostic plots. Inference on the change-point and magnitude of change was based on 95% highest posterior density credible intervals.^28^ We used R (v 4.5.0) for all analyses, and the code is available at https://github.com/yonwol3/ra-biomarker

## Results

Table 1 summarizes demographic characteristics of participants with and without clinical RA. Sex, race, and age at diagnosis were balanced between RA and non-RA groups. Among individuals who developed RA, the median age at diagnosis was 36.2 years for Cohort A and 34.7 years for Cohort B, younger than the median of 58 years reported in an early RA cohort.^29^

Figures 1 and 2 show the temporal trends in log serum biomarker levels among cases and controls. Figure 1 displays scatterplots with smoothed averages for six biomarkers tested in Cohort A, showing that five markers increase before diagnosis. The exception is RF-IgG, whose trajectory appears to change only after diagnosis. Figure 2 shows scatterplots for the eight biomarkers tested in Cohort B, where some biomarkers change discernibly before diagnosis among RA cases. For the four IgA biomarkers, the change before diagnosis is more subtle. In both figures, the post-change trends are approximately linear even though the smoothing spline could accommodate more complex nonlinear shapes, supporting our proposed segmented regression model with unknown change-points.

**Figure 1.**
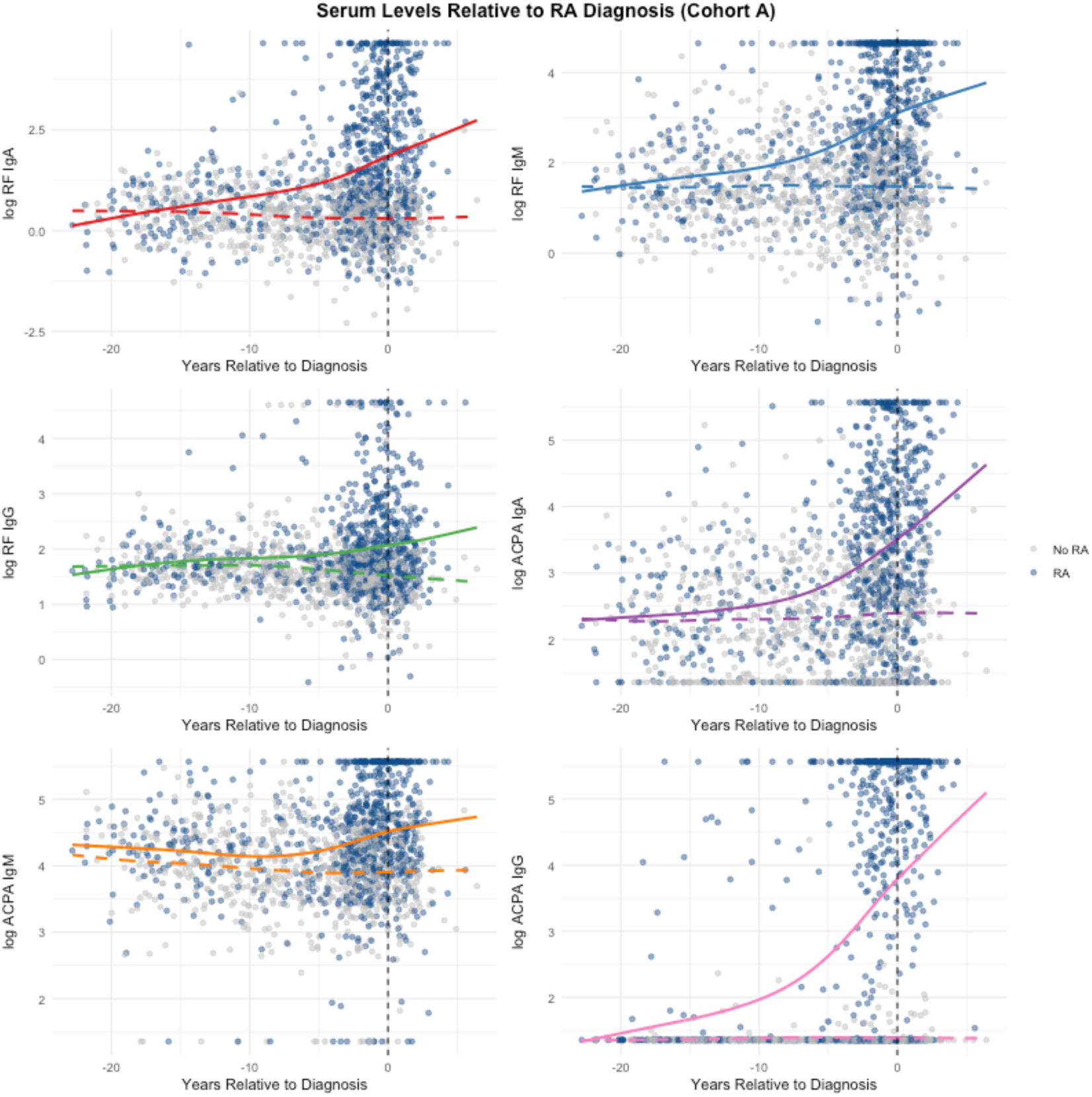
Scatterplots of the log serum antibody levels for six biomarkers (RF-IgA, RF-IgM, RF-IgG, ACPA-IgA, ACPA-IgM, and ACPA-IgG) plotted against time (in years) relative to rheumatoid arthritis (RA) diagnosis. Trends for cases (solid curves) and controls (dashed curves) are shown. Time = 0 (vertical dashed line) indicates the time of diagnosis of RA in the cases. Each plot is overlaid with a smoothing spline (4 degrees of freedom) to illustrate temporal trends in biomarker levels. Note the apparent rise in concentration for five of the six biomarkers in cases (solid lines) prior to the time of diagnosis (t=0), except for RF-IgG.

**Figure 2.**
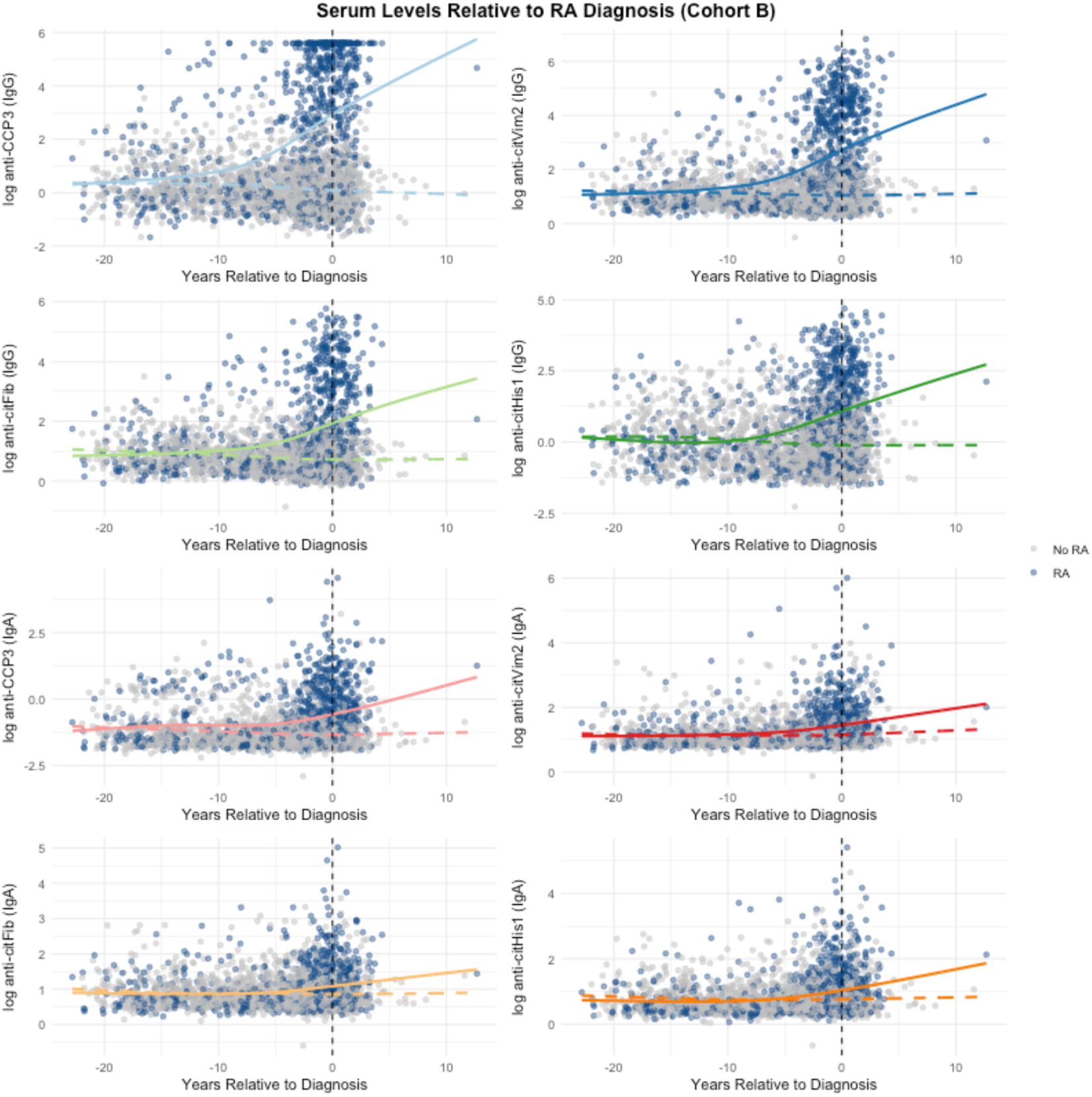
Scatterplots of the log serum antibody levels for eight biomarkers (both IgG and IgA variants of anti-CCP3, anti-citHis1, anti-citVim2, and anti-citFib) plotted against time (in years) relative to rheumatoid arthritis (RA) diagnosis. Trends for cases (solid curves) and controls (dashed curves) are shown. Time = 0 (vertical dashed line) indicates the time of diagnosis of RA in the cases. Each plot is overlaid with a smoothing spline (4 degrees of freedom) to illustrate temporal trends in biomarker levels. Observe that discernible pre-diagnostic increases in concentration are primarily visible among the IgG isotypes for cases (solid lines).

Figure 3A presents the posterior probability density functions of the change-point parameters for the six biomarkers in Cohort A. Five biomarkers (RF-IgA, RF-IgM, ACPA-IgG, ACPA-IgA, and ACPA-IgM) exhibit pre-diagnosis shifts to the initial trajectories of the RA biomarker concentrations (Table 2). RF-IgA has the earliest posterior mean change-point (9.2 years before diagnosis), followed by RF-IgM (8.2 years), ACPA-IgG (7.8 years), ACPA-IgA (6.1 years), and ACPA-IgM (5.2 years). In contrast, RF-IgG has a posterior mean change-point 4.0 years after diagnosis, near the end of post-diagnosis follow-up. None of the 95% credible intervals for magnitude of change include zero (Table 2), indicating that all biomarkers in Cohort A show changes over time among RA cases. Ranked by composite time, ACPA-IgG is first (6.6 years before diagnosis), followed by RF-IgM (6.4 years) and RF-IgA (6.0 years).

**Figure 3.**
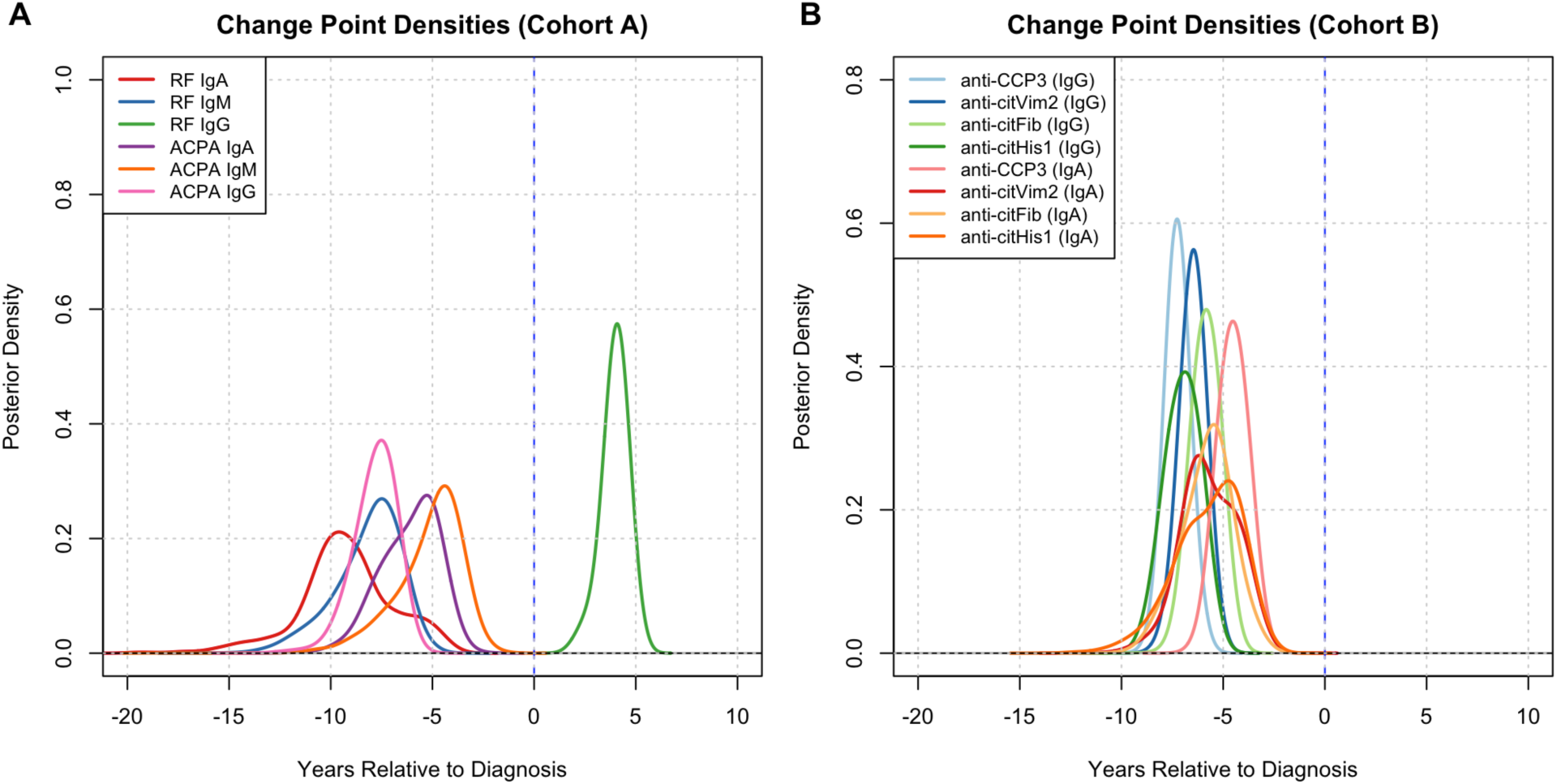
Posterior probability density functions of the change-point parameter for each biomarker. The x-axis represents years relative to RA diagnosis, with negative values indicating the pre-diagnostic period. Peaks to the left of zero indicate a high posterior probability of a pre-diagnostic change-point. **Left Panel A** depicts posterior change-point density for the six biomarkers in Cohort A (labels in the top-left corner), while **Right Panel B** depicts posterior change-point density for the eight biomarkers in Cohort B (labels in the top-right corner). The peak of each density plot corresponds to the most probable change-point estimate. RF=rheumatoid factor; Ig=immunoglobulin; ACPA=anti-citrullinated protein antibody; CCP3=cyclic citrullinated peptide-3; cit=citrullinated; His=histone; Vim=vimentin; Fib=fibrinogen.

Figure 3B presents the estimated change-points for the eight biomarkers in Cohort B. All eight biomarkers have 95% HPDIs entirely before diagnosis. Anti-CCP3 (IgG) exhibits the earliest change (7.2 years before diagnosis), followed by anti-citHis1 (IgG) (7.0 years), anti-citVim2 (IgG) (6.5 years), and anti-citFib (IgG) (5.9 years). Within each antigen, the IgG isotype has the earlier and better-identified change-point; the IgA isotypes change later (posterior means between 4.5 and 5.8 years before diagnosis) and with smaller magnitudes (Table 2). Figures S1 and S2 of the Supplement show the posterior correlations among the biomarker-specific change-points, which are moderate to strong in Cohort B.

Combining change-point timing and magnitude through the composite time *τ_k_* (defined in Methods) yields the ranking in Table 2. Because every magnitude interval excludes zero, the composite time is close to the time by which the change-point has occurred with 90% posterior probability, so the ranking depends on the precision of a change-point as well as its location. In Cohort A, RF-IgA has the earliest posterior mean change-point but ranks third on *τ_k_*, because its change-point interval is the widest; ACPA-IgG, with a narrower change-point interval, ranks first on *τ_k_* (Table 2). In Cohort B, the rankings by composite time and by average change-point are nearly identical, and all four IgG isotypes precede all four IgA isotypes on both.

The sensitivity analysis (Supplement) fixes the residual correlation to the identity. The two fits agree on which biomarkers change before diagnosis and on the composite-time ordering of IgG before IgA in Cohort B. They differ where the data are least informative. Without the residual correlation, the change-point for RF-IgG in Cohort A is essentially unidentified: its 95% HPDI runs from −16.5 to 5.1 years, against 2.6 to 4.9 years under the primary model. RF-IgA moves from third to first on composite time. Three of the four IgA intervals in Cohort B also widen.

## Discussion

Our model estimates within-case acceleration in autoantibody concentrations rather than divergence between cases and controls, and reports posterior uncertainty in both the timing and magnitude of change. The model also allows for comparison of change-points across autoantibody types (including isotypes, e.g., IgA *vs.* IgG), which may help disentangle the distinct biological processes driving disease evolution at different stages and inform future prediction of the onset of clinical RA.^30^

Our findings for Cohort A complement those of Kelmenson and colleagues,^14^ who analyzed the same cohort. While this previous article identified the earliest time point of statistical divergence between cases and controls, which indicates when early RA-related autoimmunity develops years before arthritis is identified, our model identifies the point of trajectory acceleration within cases, a measure that reflects underlying pathogenic shifts. For example, we identified the change-point for ACPA-IgG at 7.8 years before diagnosis, suggesting an increase in the rate of autoantibody production at this time. This can be reconciled with the earlier divergence reported approximately 17.9 years before diagnosis, which may represent the initial, slow-rising phase of autoimmunity that precedes this acceleration.^14^

Within Cohort A, RF-IgA had the earliest posterior mean change-point, but its wide change-point interval placed it third under the composite ranking, behind ACPA-IgG and RF-IgM. The composite time should therefore be read together with the underlying ***δ*** and ***γ*** estimates. In contrast, in Cohort B, all eight biomarkers changed before diagnosis, with the IgG responses changing earlier and more steeply than the IgA responses. The residual correlation narrowed three of the four IgA intervals (Supplement). The ordering of change-points across citrullinated targets during the preclinical phase of RA is in line with previous studies and the concept of B-cell diversification and epitope spreading.^31–33^

IgA autoantibodies in the serum can be markers of mucosal processes, and mucosal interfaces are hypothesized to be early sites of RA autoimmunity.^34^ Because in both cohorts the IgG responses to citrullinated antigens changed earlier than the corresponding IgA responses, systemic IgG responses may be the earlier and stronger serum signal, with the serum IgA signal present but later and weaker. This may indicate that IgA autoantibodies (and potentially mucosal processes) are important in the final transition from a preclinical state to clinical RA.

However, the IgA autoantibodies measured in the two cohorts differed (e.g., RF-IgA in Cohort A *vs.* anti-citFib IgA in Cohort B), as did the assay platforms (ELISA vs. particle-based multianalyte technology) and the matching ratios (1:1 vs. 1:2). Where the panels overlap, ACPA-IgG in Cohort A and anti-CCP3 (IgG) in Cohort B had nearly identical composite times (6.6 and 6.7 years before diagnosis), and the change-point intervals for ACPA-IgA and anti-CCP3 (IgA) overlapped (Table 2). Although ACPA-IgG ranked first on composite time in Cohort A, RF-IgA had the earliest average change-point, ahead of both IgG assays. This suggests that certain IgA autoantibodies may occur earlier, and that there may be subsets of individuals (e.g., endotypes) where IgA responses are earlier than IgG. The RF-IgA change-point was also the least precise in Cohort A, which could reflect greater assay variability or a rise confined to a subset of cases. The model assumes one change-point shared by all cases and cannot separate these explanations. Mucosal processes may also occur that are unmeasured in blood. These issues will need investigation in future studies.

Lastly, in Cohort A, RF-IgG shifted only after diagnosis, possibly reflecting post-clinical biology or treatment effects; this change-point falls near the end of follow-up, when few samples were collected, and warrants further investigation.

The matched case-control design supports adjustment for confounding by the matching factors. Limitations include reliance on known RA cases, which may limit prospective applicability to populations with abnormal biomarkers but uncertain progression to clinical disease, and a likely true data-generating process with random slopes, individual-level change-points, or higher-order polynomial terms not captured by the current specification. Removing the residual correlation widens the intervals for the weakly identified biomarkers but leaves the set of pre-diagnostic biomarkers unchanged (Supplement). The pair-level random intercept in Cohort B further shares a baseline between one case and two controls, which does not fully distinguish between-individual variation among controls within a matched set; a nested individual-level effect could address this but may introduce identifiability challenges given the sparse within-individual observations.

Autoantibodies are detectable years before clinical RA, yet it remains unclear which autoantibody-positive individuals will progress and when to intervene.^3^ The change-points estimated here indicate when each autoantibody begins to accelerate among individuals who later developed RA, information that a single positive or negative result does not provide. In autoantibody-positive individuals followed with serial quantitative testing, such an acceleration, or a later rise in IgA isotypes, could help identify those approaching clinical RA and the time to offer preventive treatment, which prospective studies will need to confirm. The Bayesian multivariate segmented regression described here estimates change-points and their magnitudes in correlated longitudinal biomarkers and can be applied beyond preclinical RA.

## Ethics approval

Use of anonymized clinical data and biospecimens was approved by Institutional and Regulatory Review Boards at the University of Colorado (IRB #02-297), Walter Reed National Military Medical Center (IRB #20293), and the U.S. Army Medical Research and Development Command, Office of Research Protections (ORP), Human Research Protection Office (HRPO) (HRPO Log Number E01484.1a).

## Author contributions

Y.F.W. and K.P.J. conceived and designed the analytical strategy, developed the statistical methodology, performed the data cleaning and analysis, interpreted the results, and drafted the manuscript. K.D.D. provided the data, contributed to the interpretation of results, and critically revised the manuscript. J.D.E. provided the data and critically revised the manuscript. B.D.W. contributed to the interpretation of results and critically revised the manuscript. A.M.J., M.L.F., and M.M. critically revised the manuscript for important intellectual content. K.P.J. is the guarantor for this paper.

## Supplementary data

Supplementary data are available at *JCE* online.

## Data availability

Because of regulatory issues, the data from this study are not available. However, code to implement this methodology can be found here: https://github.com/yonwol3/ra-biomarker

