## supplemental_materials for "Detecting change-points in preclinical rheumatoid arthritis biomarkers using Bayesian multivariate segmented regression"

The main text presents the primary analysis, which treats values at the upper limit of detection as right-censored and models an unstructured residual correlation among biomarkers. This supplement gives the prior specifications, records the computational details of the data augmentation, and reports a sensitivity analysis that fixes the residual correlation to the identity, leaving every other model component unchanged.

**Prior Distributions**

We suppose the slope parameters $\boldsymbol{\beta}_1$, $\boldsymbol{\beta}_2$, and $\boldsymbol{\gamma}$ have a multivariate normal distribution, with mean zero and a diagonal covariance matrix with diagonal elements equal to 4 on the log scale. The intercept vector $\boldsymbol{\theta}$ has a multivariate normal prior with mean zero and a diagonal covariance matrix with diagonal elements equal to $10^6$ for Cohort A and $10^8$ for Cohort B. For $k = 1,2,\dots,K$, the variance components $\sigma_k$ and $\omega_k$ both have a half-Cauchy(0, 5) prior.[1] Change-points $\delta_k$ have a continuous uniform prior over [−20, 10] years.[2] The correlation matrix $\boldsymbol{\Phi}$ has an LKJ(2) prior on its Cholesky factor.[3,4] The sensitivity model uses the same priors with the correlation matrix fixed to the identity. All choices are non- or weakly informative.

**Modeling Details**

Both models are fit to log-transformed concentrations. For a participant $i = 1,2,\dots,n$, with measurement $j = 1,2,\dots,m_i$, and for biomarker $k = 1,2,\dots,K$ ($K = 6$ for Cohort A; $K = 8$ for Cohort B), we denote the outcome vector as $\boldsymbol{Y}_{ij} = \left[Y_{ij1}, Y_{ij2}, \dots, Y_{ijK}\right]^{\mathrm{T}}$ , with each element the log concentration of the corresponding biomarker. The response distribution follows

$$\boldsymbol{Y}_{ij} \sim \text{Normal}(\boldsymbol{\mu}_{ij}, \boldsymbol{\Sigma}).$$

If a biomarker concentration $Y_{ijk}$ equals its maximum detectable value, $U_k$, the value is treated as right-censored. Censoring is handled by data augmentation.[5,6] Each censored cell receives a latent exceedance constrained to be non-negative, and the completed value equals the detection limit plus the exceedance. The completed outcome vectors follow the multivariate normal likelihood, evaluated through the Cholesky factor of the residual covariance, and the latent exceedances are sampled jointly with the model parameters in Stan. Integrating over the exceedances recovers the censored-data likelihood. The random intercepts use a non-centered parametrization to improve mixing. In the sensitivity model, we fix the residual correlation to the identity, so the residual covariance reduces to $\boldsymbol{\Sigma} = \text{diag}(\boldsymbol{\sigma}^2)$, the likelihood factorizes across biomarkers, and each censored contribution is evaluated analytically with the upper-tail normal probability, with a small numerical floor to avoid log(0); no latent values are required. No prior is then needed for the correlation matrix $\boldsymbol{\Phi}$. The random intercept $\boldsymbol{\alpha}_{l[i]} \sim \text{N}(\boldsymbol{\theta}, \boldsymbol{\Omega})$ is defined just as it is in the main text, and the mean model remains unchanged.

Both models share the priors listed above and differ only in the residual correlation structure. The sensitivity results are summarized in Table S1, with biomarkers ranked by the earliest composite time (defined in the main text).

**Sensitivity Analysis Results**

For Cohort A, the diagonal model identifies the same five pre-diagnostic biomarkers as the primary model, and the point estimates shift earlier: RF IgA moves from -9.2 to -10.6 years and ACPA IgM from -5.2 to -6.1 years. On composite time, RF IgA ranks first under the diagonal model, ahead of RF IgM and ACPA IgG (Table S1). The clearest difference is RF IgG. Under the primary model its change-point is identified after diagnosis (4.0 years; 95% HPDI: 2.6, 4.9); under the diagonal model the interval spans -16.5 to 5.1 years (Table S1). This change-point is identified only when the residual correlation is modeled. Figure S3 shows the change-point posterior densities for Cohort A biomarkers, while Figure S4 shows the change-point posterior densities for Cohort B biomarkers. The correlated and diagonal models give broadly similar posterior densities for the well-identified biomarkers.

For Cohort B, all eight biomarkers retain pre-diagnostic change-points and composite times under the diagonal model, and the IgG isotypes again precede the IgA isotypes on composite time, with anti-CCP3 (IgG) earliest (Table S1). The intervals for three of the four IgA isotypes are wider than under the primary model. The agreement between the two fits supports the Cohort B ranking.

**Table S1:** Summaries of posterior output from the sensitivity model with a diagonal residual covariance, for Cohort A and Cohort B (compare Table 2).

| BIOMARKER | COMPOSITE TIME (YEARS) | AVERAGE CHANGE-POINT [95% HPDI] | MAGNITUDE OF CHANGE MEAN [95% HPDI] |
|---|---|---|---|
| **COHORT A** | | | |
| RF IgA | -8.1 | -10.6 [-15.8, -6.7] | 0.14 [0.10, 0.18] |
| RF IgM | -7.1 | -8.9 [-12.2, -6.1] | 0.17 [0.12, 0.22] |
| ACPA IgG | -7.0 | -8.1 [-9.7, -6.5] | 0.27 [0.22, 0.32] |
| ACPA IgA | -5.1 | -6.6 [-8.8, -4.4] | 0.15 [0.11, 0.19] |
| ACPA IgM | -4.3 | -6.1 [-9.0, -3.5] | 0.08 [0.05, 0.11] |
| RF IgG | 4.1 | 0.3 [-16.5, 5.1] | 1.38 [0.02, 3.26] |
| **COHORT B** | | | |
| anti-CCP3 (IgG) | -7.0 | -7.7 [-8.8, -6.6] | 0.34 [0.29, 0.38] |
| anti-citHis1 (IgG) | -6.5 | -7.6 [-9.3, -5.8] | 0.17 [0.14, 0.20] |
| anti-citVim2 (IgG) | -6.2 | -6.9 [-8.1, -5.8] | 0.22 [0.19, 0.25] |
| anti-citFib (IgG) | -5.4 | -6.3 [-7.5, -5.0] | 0.18 [0.15, 0.21] |
| anti-citVim2 (IgA) | -4.8 | -6.5 [-9.6, -3.6] | 0.04 [0.03, 0.06] |
| anti-citHis1 (IgA) | -4.8 | -6.6 [-9.8, -3.7] | 0.05 [0.03, 0.07] |
| anti-citFib (IgA) | -4.7 | -6.5 [-9.7, -3.7] | 0.04 [0.02, 0.06] |
| anti-CCP3 (IgA) | -4.1 | -5.1 [-6.6, -3.6] | 0.12 [0.09, 0.14] |

Magnitudes of change are slopes of log concentration per year. **Abbreviations**: RF=rheumatoid factor; Ig=immunoglobulin; ACPA=anti-citrullinated protein antibody; CCP3=cyclic citrullinated peptide-3; cit=citrullinated; His=histone; Vim=vimentin; Fib=fibrinogen; HPDI=highest posterior density interval.

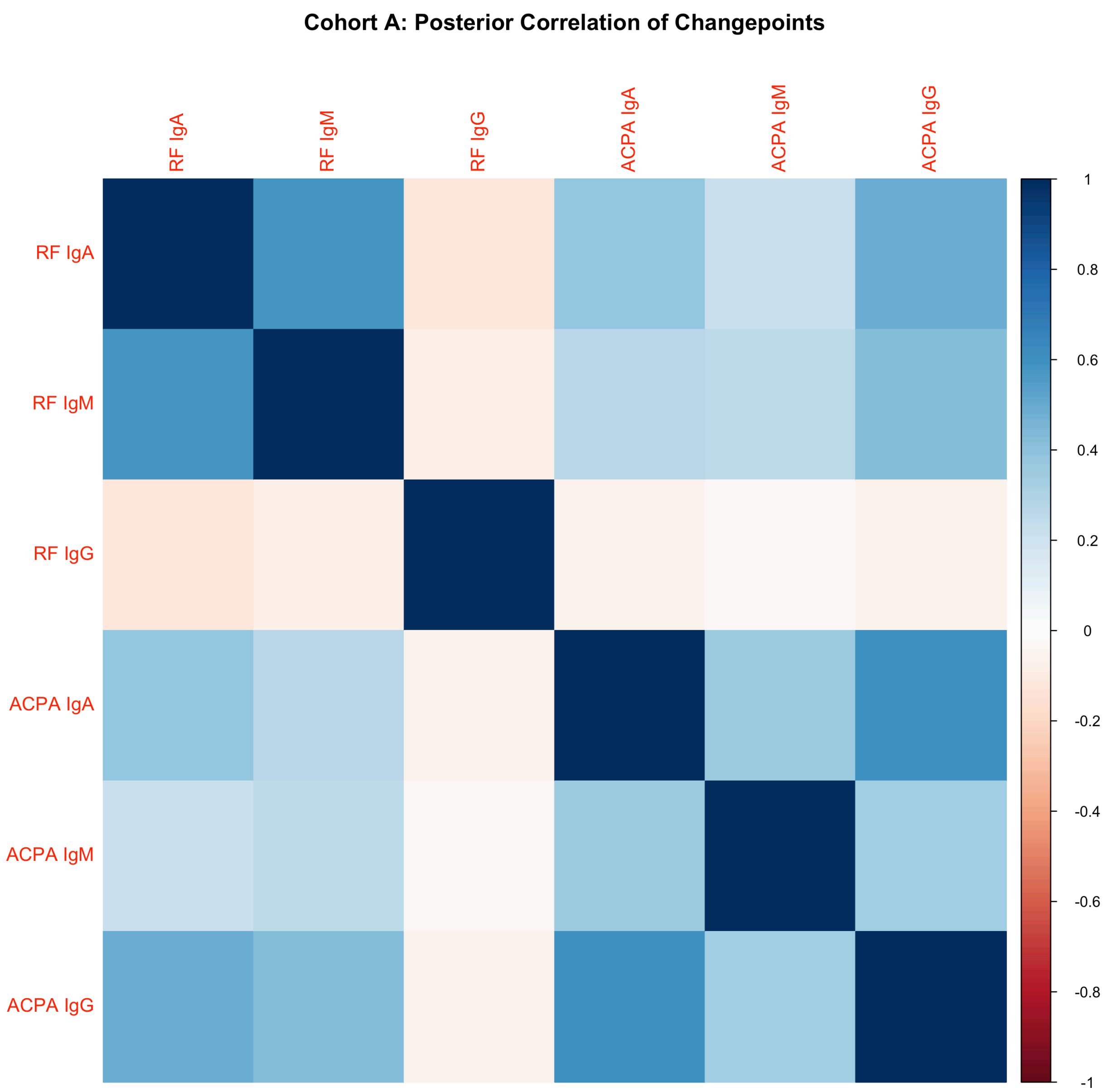


**Figure S1.** Posterior correlation matrix of the change-point parameters for the six biomarkers in Cohort A under the primary censored model with unstructured residual correlation. Each cell displays the correlation between two biomarker-specific change-point parameters across MCMC draws, with color intensity indicating the strength and direction of correlation.

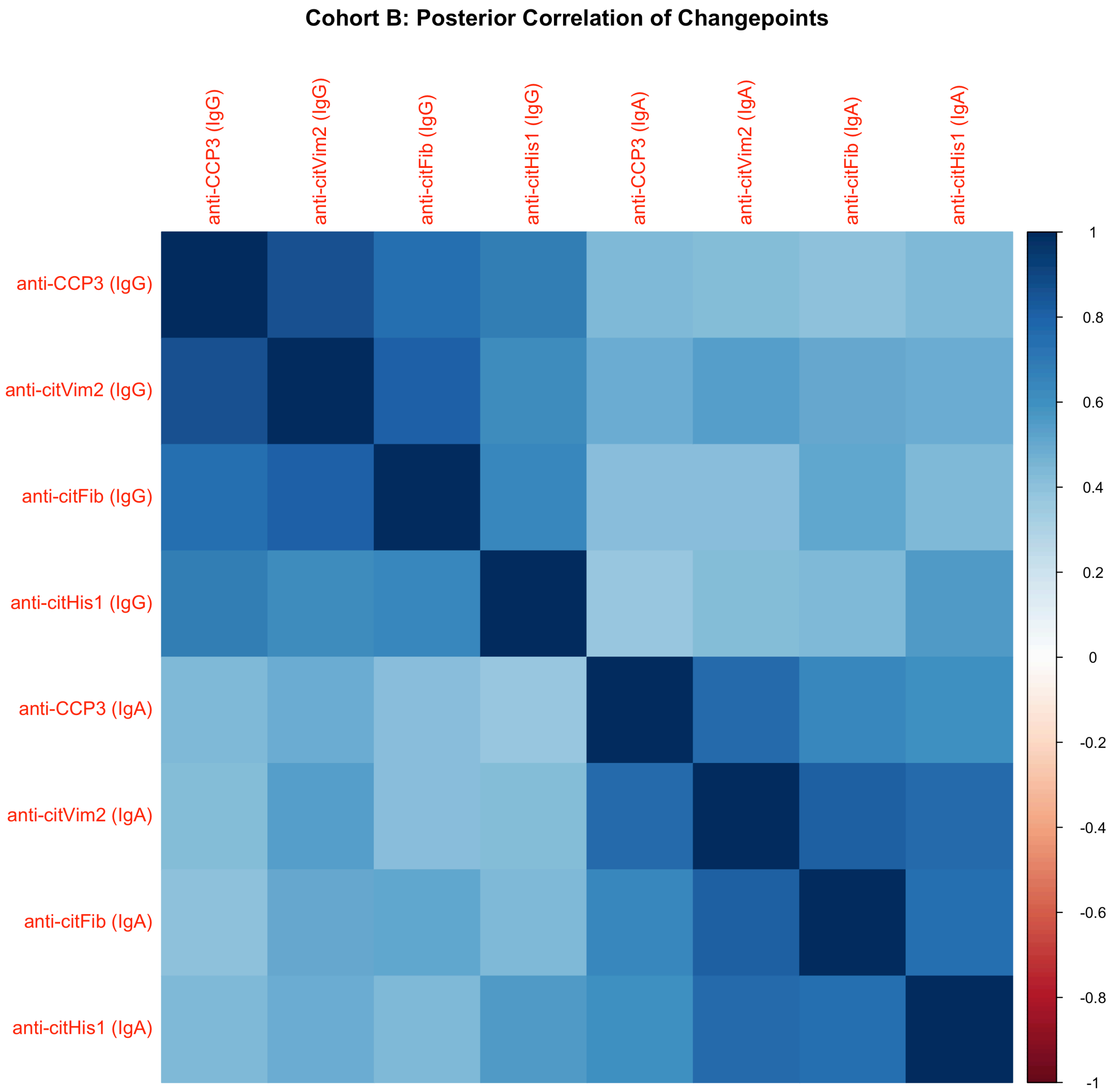


**Figure S2.** Posterior correlation matrix of the change-point parameters for the eight biomarkers in Cohort B under the primary censored model with unstructured residual correlation. Each cell displays the correlation between two biomarker-specific change-point parameters across MCMC draws, with color intensity indicating the strength and direction of correlation.

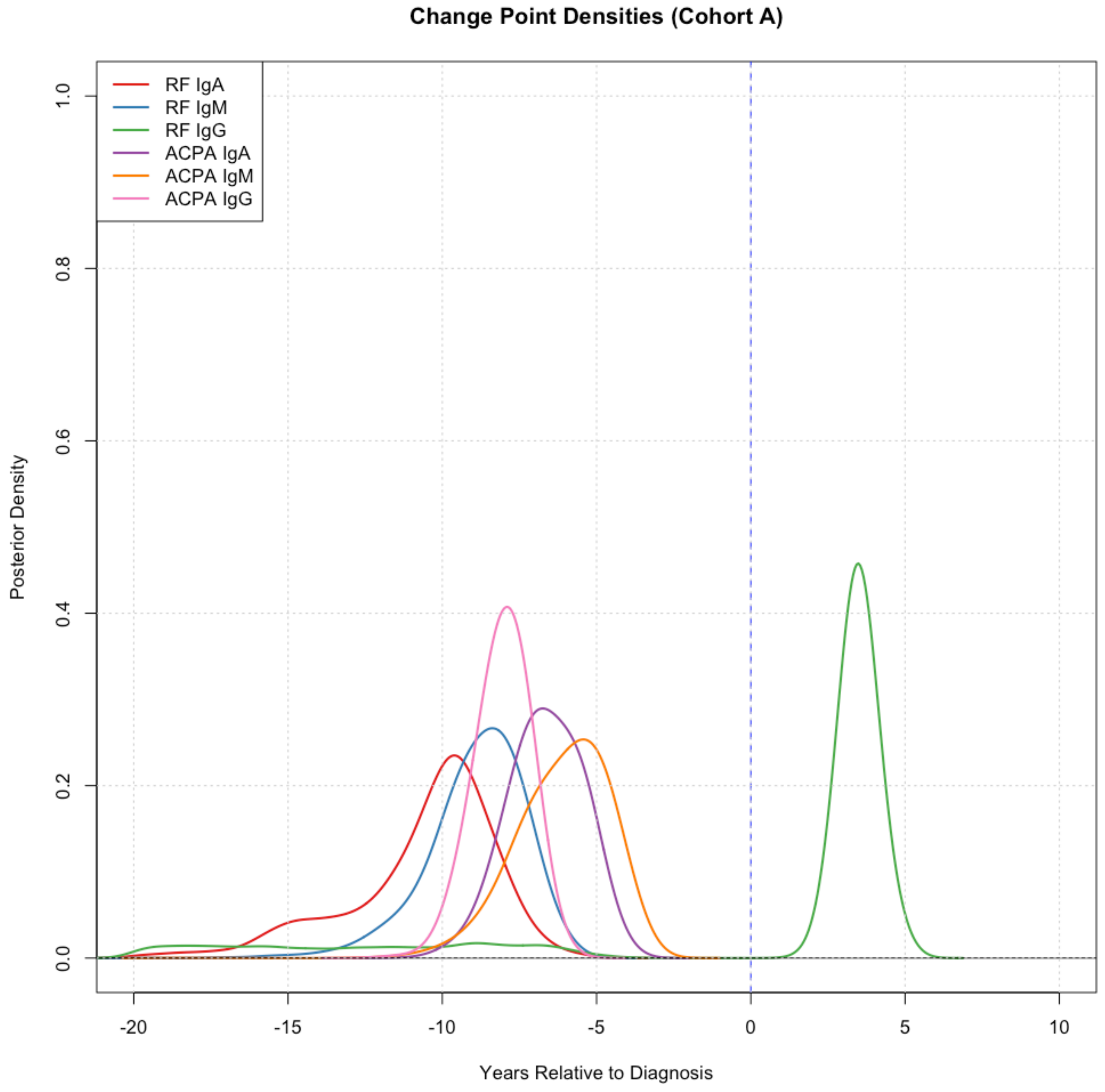


**Figure S3.** Posterior probability density functions of the change-point parameter for each biomarker from the sensitivity (diagonal covariance) model. The six biomarkers in Cohort A are labeled in the top-left corner. The peak of each density plot corresponds to the most probable change-point estimate.

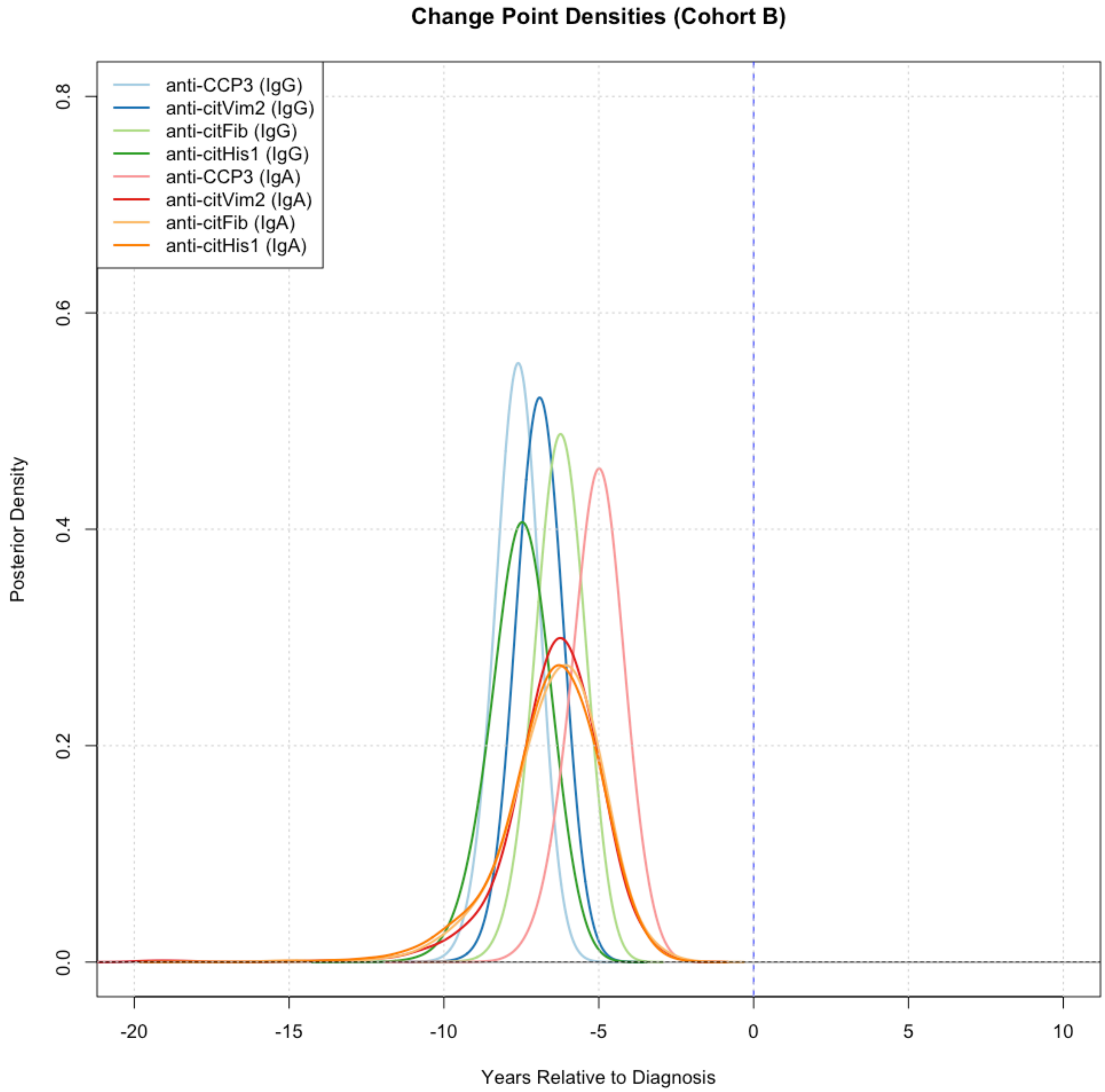


**Figure S4.** Posterior probability density functions of the change-point parameter for each biomarker from the sensitivity (diagonal covariance) model. The eight biomarkers in Cohort B are labeled in the top-left corner. The peak of each density plot corresponds to the most probable change-point estimate.
